# Comparison of quality of sepsis care among patients with vs. without acute mental health crises

**DOI:** 10.64898/2026.02.09.26345933

**Authors:** Rida Nasir, Yi-Ru Chen, Melva Morales Sierra, Jaime Jacob, Lisa Iyeke, Lindsay Jordan, Khatija Paperwalla, Mark Richman

## Abstract

**Introduction:** Sepsis is a life-threatening ailment caused by an exaggerated immune response to infection that poses a major health problem, with increasing prevalence, high costs, and poor outcomes. Improved outcomes are seen in patients when providers follow the Surviving Sepsis Campaign recommended clinical practice guidelines for identifying and treating sepsis using a 3-hour and 6-hour bundle after sepsis is suspected. Previous research has shown patients with mental health issues receive worse quality of diabetes and cardiac care and have poorer outcomes compared with those without mental health issues. Similarly, patients with mental health issues may receive worse sepsis care due to inability to explain symptoms, agitation, etc. This study explores sepsis quality of care among patients with vs. without an acute mental health crisis, and whether patients with certain mental health issues were more likely to receive sepsis bundle care than others.

**Methods:** Using data extracted from 2018-2019 at the Long Island Jewish Medical Center Emergency Department (ED), patients who met sepsis inclusion criteria were grouped into either having, or not having, a severe mental illness crisis on the basis of whether physical or chemical restraints were used in the ED. Patients with a history of a severe mental illness, but who were not in a severe mental health crisis, were grouped with the patients without mental health illness, as, in the absence of an acute psychiatric problem, their mental health issue unlikely affected sepsis care. We describe demographic characteristics of both groups and performed a univariate analysis using Student’s T-test to compare the percent of those with vs. without acute mental health crisis who received full 3- and 6-hour sepsis bundle care. Patients with an acute mental health crisis were grouped according to “cognitive” (eg, dementia) vs. “non-cognitive” (eg, schizophrenia) disorders.

**Results:** Comparing those with vs. without acute mental health crisis, there was no difference in the percent of patients who received 3-hour sepsis bundle care (80.7% vs 74.9%, p = 0.1456). However, among patients who received the 3-hour bundle, a significantly-greater percent of those with an acute mental health crisis received the 6-hour sepsis bundle (51.0% vs. 30.7%, p <0.0001). There was no difference between different groups of patients with mental health issues (eg, “cognitive” vs. “non-cognitive”) with respect to receiving 3- or 6-hour sepsis bundle care.

**Discussion:** Surprisingly, although there was no significant difference in likelihood to receive a 3-hour sepsis bundle among patients with vs. without an acute mental health crisis, those with an acute mental health crisis were more-likely to receive 6-hour care. We suspect this difference might be due to increased attention paid to patients with an acute mental health crisis, including more-frequent room visits by hospital staff or more concerns among family members. No particular set of mental health conditions was associated with receiving or not receiving appropriate care. Future research could address possible confounding factors, go into more detail about the specific component of the sepsis protocol that patients failed to receive, and specify what aspects of a mental health crisis affected treatment plans. Future studies are needed to assess possible associations between severe mental illness crisis, bundle care, and mortality in relation to ED, Intensive Care Unit (ICU), or hospital length-of-stay (LOS).

## Background

Sepsis is a life-threatening ailment caused by an exaggerated immune response to infection that results from the body’s response to infection, which causes injury to its own tissues and organs. Sepsis is a major health problem with increasing prevalence, high costs, and poor outcomes. In the United States, sepsis accounts for about 5.2% of hospital expenditures, more than $20 billion per year, and hospitalizations for more than 1 million people.^1^ Severe sepsis and septic shock are a subset of sepsis conditions.

This subset of conditions accounts for 25–50% of in-hospital mortality, ranging from 45% to 65% after six months of discharge.^2^ Mortality rates rise with sepsis severity: 10-20% for sepsis, 20-40% severe sepsis, and 40-80% for septic shock.^3^ If patients with these conditions survive, they have increasing chronic complications, morbidity, high costs of care, and decreasing quality of life.^4,5^ Many of the over 1.5 million annual cases of sepsis in the United States present to the Emergency Department (ED). ^6,7^ Improved outcomes, including decreased mortality, are achieved with prompt identification of sepsis and treatment with broad-spectrum antibiotic agents and intravenous fluids,^8^ as recommended in the Surviving Sepsis Campaign and endorsed by clinical practice guidelines and the Centers for Medicare and Medicaid Services (CMS). ^9,10^

Per the Surviving Sepsis Campaign, sepsis is defined as having ≥2 SIRS criteria (temperature >38.0°C or hypothermia <36.0°C, heart rate >90 beats/minute, respiratory rate >20 breaths/minute, leukocytosis >12*10^9^/L or leukopenia <4*10^9^/L) and a likely bacterial source of their SIRS. Patients meeting these criteria should, within 3 hours of sepsis being suspected, have: 1) Serum lactate measured, 2) For patients with elevated lactate or hypotension (mean arterial pressure (MAP <65 mmHg), intravenous fluid (IVF) bolus of at least 30 mL/kg administered, unless contraindicated by evidence or risk of volume overload, 3) In fluid-non-responsive patients with elevated lactate or hypotension, giving vasopressors to raise blood pressure, 4) Blood cultures drawn before antibiotics given, 5) Broad-spectrum antibiotics administered after blood cultures drawn, 6) For patients with elevated serum lactate, a repeat serum lactate and re-assessment of the need for additional IVF within 6 hours of sepsis being suspected, to determine whether tissue perfusion improved with initial IVF and vasopressors.^10^

Patients may fail to receive the complete bundle for a variety of reasons. There may be a delay in recognizing sepsis. Patients may have a contraindication to the full 30 mK/kg IVF bolus, or an advanced directive limiting use of bundle interventions. They might: 1) be currently psychotic or manic and unable to explain their symptoms appropriately, 2) not understand their condition or the care they’re offered, 2) refuse care, 3) be agitated or aggressive, or 4) take medications whose side effects could be misinterpreted as sepsis (eg, neuroleptic malignant syndrome (NMS)).

Previous research has demonstrated patients with severe mental illness and cardiovascular disease or diabetes are 30% less likely to receive recommended cardiovascular or diabetes care, including hospitalization, diagnostic tests, appropriate medications, and invasive procedures.^11^ It is possible such disparities apply to severe sepsis or septic shock, as well. However, very few such studies have been performed. A national cross-sectional study in Sweden found patients with severe mental health disorders had increased risk of death associated with influenza/pneumonia (OR = 2.06, 95% CI [1.87–2.27]) and sepsis (OR = 1.61, 95% CI [1.38–1.89]). They also had an increased risk of hospitalization associated with influenza/pneumonia (OR = 2.12, 95% CI [2.03–2.20]) and sepsis (OR = 1.89, 95% CI [1.75–2.03]).^12^

The primary aim of this exploratory study was to determine whether having a severe, concurrent mental illness crisis may impact the quality and timeliness of sepsis bundle care provided to patients, as indicated by the percent of patients in each group who received 3- and 6-hour bundle-compliant care. We hypothesized that patients with a concurrent severe mental illness crisis will have had increased risk of not receiving the full CMS sepsis bundle compared with patients without severe, concurrent mental illness. If such a disparity is identified, future studies will investigate confounding variables that might explain such variation in quality of care. The secondary aim was to determine which psychiatric illness crises confer the most risk for not receiving the full bundle.

## Methods

Northwell Health is a 23-hospital health system largely operating in Long Island and New York City. Long Island Jewish (LIJ) Medical Center is a 583-bed tertiary care teaching hospital serving an ethnically and socio-economically diverse population. The adult ED sees approximately 100,000 patients per year. Members of LIJ’s Department of Quality Services collect and submit sepsis data to state and federal agencies.

Patients with a contraindication to bundle components or an advanced directive are removed from the sepsis database. We extracted data from LIJ from 2018 and 2019.

### Inclusion criteria

1. Location: LIJ ED
2. Dates of patient ED presentation: 1/1/18-12/31/19
3. Sepsis (per LIJ sepsis data extraction and reporting management system)

### Exclusion criteria

1. Pregnant
2. Patients with advanced directives for non-aggressive medical care (these will already have been excluded through the sepsis data extraction and reporting management system)

Patients who meet inclusion criteria will be stratified into either having, or not having, a severe mental illness crisis (including being severely agitated) at the time of ED presentation based on the conditions/ICD-10 codes below (**Appendix A**) plus a chart review focusing on the ED note and ED or inpatient Psychiatry consult evaluation.

Presence of severe, concurrent mental illness will be determined by whether:

1. The ED note suggests the patient has a history of dementia or mental illness and worsening agitation, or
2. The patient had a Psychiatric consult during their index (sepsis) admission stating the patient was in a mental health crisis just prior to or at the time of ED presentation (Psychiatry consult notes include collateral information from family, group homes, or psychiatric institutions reflecting the patient’s mental state and behavior prior to arrival.)

Patients with a history of a severe mental illness (eg, major depression), but who were not in a severe mental health crisis (ie, not psychotic) at the time of ED presentation will be included in the control group, as their non-severe mental illness would be unlikely to affect their sepsis care.

### Statistics

#### 1. Sample size calculation

To estimate sample size for the primary aim (% of patients with severe mental illness vs. % without severe mental illness who received full sepsis bundle care), we looked at 2019 LIJ data for all patients with severe sepsis or septic shock. Sixty-eight percent (575/847) met the 3-hour bundle (32% did not); 36% (300/834) met the 6-hour bundle (64% did not). Assuming that, similar to patients with severe mental illness and cardiovascular disease or diabetes, patients with severe mental illness are 30% less likely to receive recommended sepsis care, we would expect 52% of patients with severe mental illness not to meet the 3 hour bundle and 75% not to meet the 6-hour bundle. To achieve statistical significance with a power of 80% and a p-value for statistical significance a priori at <0.05 would require at least 65 patients per arm (severe mental illness crisis vs. no severe mental illness crisis).

#### 2. Data analysis

Student’s T-test will be used to compare proportions who received vs. didn’t receive full sepsis bundle care comparing patients with vs. without acute mental health crises.

This project was deemed to fall under the realm of performance improvement by the Institutional Review Board (IRB #: 24-0076).

## Results

Patients with a history of mental illness were significantly older than those without, and their mental illnesses were more-likely dementia and other neuro-cognitive disorders (**Table 1).** The majority of patients with mental health disorders had Medicare (**Table 2**), while patients without mental health issues were more-likely to have private insurance. Significantly-more patients with mental health disorders arrived from a psychiatric hospital or facility, while most of the patients without mental health issues came from home (**Table 3**). Patients with acute mental health crises were also significantly more-likely to have comorbidities such as (AIDS, or immune-modifying conditions) (**Table 4**). Finally, the source of sepsis among patients with mental health issues was often the respiratory system, while the source among those without mental health issues was more-often the urinary tract system (**Table 5**).

**Table 1.**

| Characteristic | History of mental health illness<br>(N = 157, 26.9% of total)) | No history of mental health illness<br>(N = 426, 73.1% of total) | P-value |
| --- | --- | --- | --- |
| Age (years) | 77.9 | 67.6 | <0.0001 |
| Gender (% male) | 49.7 | 50.1 | 0.93 |

**Table 2.**

| Insurance | History of mental illness (%) (N = 157) | No history of mental illness (%) (N = 426) | P-value |
| --- | --- | --- | --- |
| Blue Cross | 3.2 | 3.5 | 0.85 |
| Medicaid | 29.3 | 33.4 | 0.35 |
| Medicare | **58** | 47.1 | **0.02** |
| Other non-Federal Program | 0.6 | 0.5 | 0.84 |
| Other private insurance company | 7.6 | **15.2** | **0.02** |
| Self-pay | 1.3 | 0.5 | 0.33 |

**Table 3.**

| Source of arrival | % with history of mental health illness | % without history of mental crisis | P-value |
| --- | --- | --- | --- |
| Clinic | 3.2 | 8.9 | 0.02 |
| Home or assisted living facility | 56.7 | 75.2 | <0.01 |
| Hospice | 0.6 | 0.7 | 0.94 |
| Hospital | 0.6 | 1.4 | 0.45 |
| Psychiatric facility or group home | 32.5 | 2.1 | <0.01 |
| Skilled Nursing Facility | 6.5 | 11.7 | 0.07 |

**Table 4.**

| Comorbidity | % with history of mental health illness | % without history of mental illness | P-value |
| --- | --- | --- | --- |
| AIDS or HIV | 3.8 | 0.5 | <0.01 |
| Altered mental status | 70.1 | 43.2 | <0.0001 |
| Chronic liver disease | 0.6 | 2.6 | 0.13 |
| Chronic renal failure | 6.4 | 8 | 0.52 |
| Chronic respiratory failure | 0 | 0.5 | 0.39 |
| Congestive heart failure | 19.1 | 15 | 0.24 |
| Diabetes | 40.6 | 43.3 | 0.58 |
| Immune-modifying medications | 7 | 20.7 | <0.01 |
| Lower respiratory infection | 45.2 | 46.5 | 0.78 |
| Lymphoma, leukemia, or multiple myeloma | 0.6 | 5.9 | <0.01 |
| Metastatic cancer | 3.8 | 12.7 | <0.01 |
| Organ transplant | 0.6 | 2.1 | 0.21 |
| ≥1 comorbidity | 72.6 | 60.3 | <0.01 |

**Table 5.**
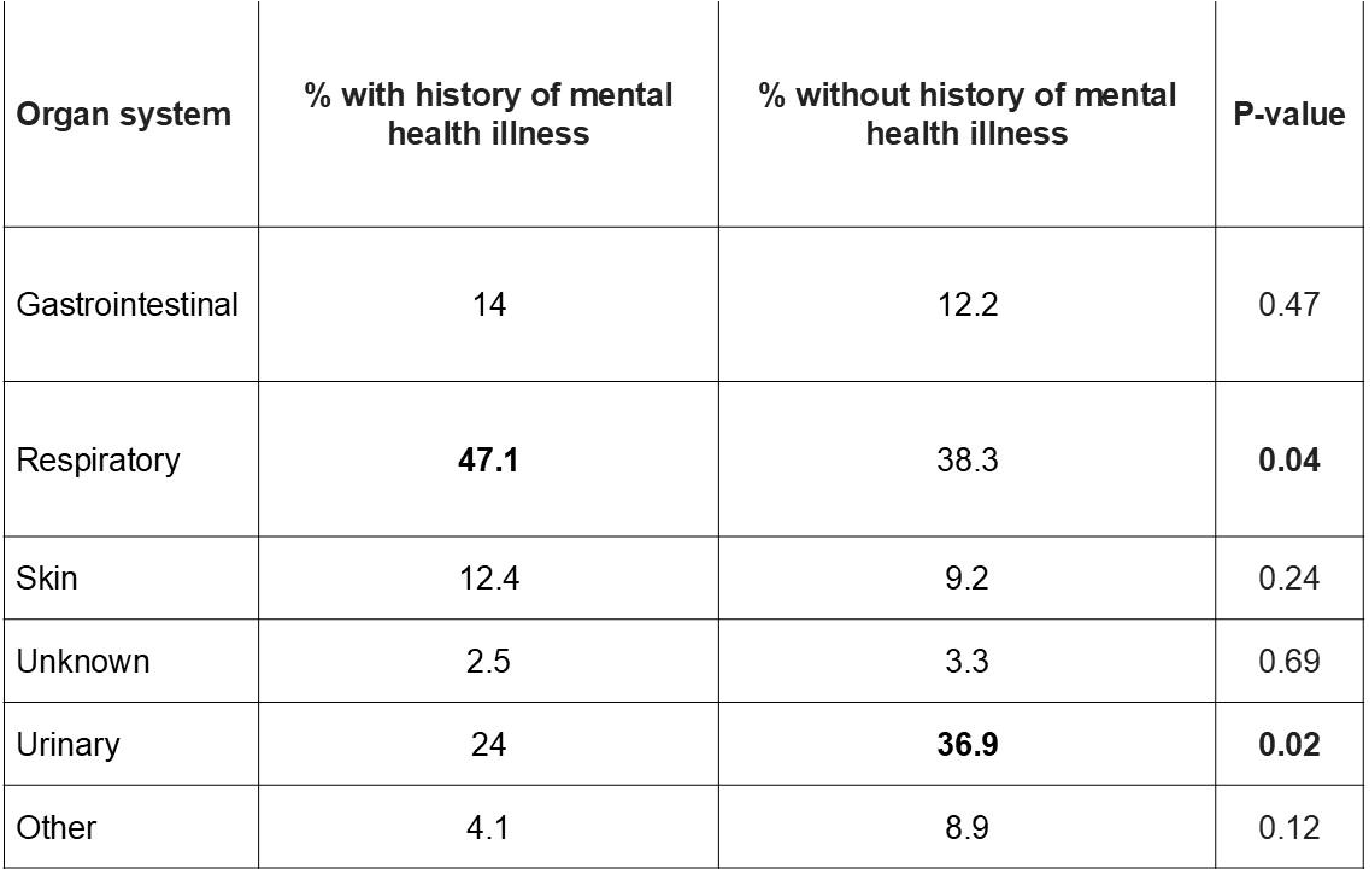

| Organ system | % with history of mental health illness | % without history of mental health illness | P-value |
| --- | --- | --- | --- |
| Gastrointestinal | 14 | 12.2 | 0.47 |
| Respiratory | **47.1** | 38.3 | **0.04** |
| Skin | 12.4 | 9.2 | 0.24 |
| Unknown | 2.5 | 3.3 | 0.69 |
| Urinary | 24 | **36.9** | **0.02** |
| Other | 4.1 | 8.9 | 0.12 |

There were no statistically-significant differences in the percent of patients with vs. without an acute mental health crisis who received 3-hour bundle-compliant care (p = 0.1456). However, a greater percent of patients with an acute mental health crisis received 6-hour bundle care (p = 0.0001) (**Table 6, Figure 1**)) That is, among those with initially-elevated serum lactate, a repeat serum lactate and re-assessment of the need for additional IVF within 6 hours of sepsis being suspected was performed more-often for patients with vs. without an acute mental health crisis.

**Table 6.**

| Received full sepsis bundle care | % with mental health crisis | % without mental health crisis | P-value |
| --- | --- | --- | --- |
| 3-hour bundle | 80.7 | 74.9 | 0.1456 |
| 3- + 6-hour bundle | 51.0 | 30.7 | **0.0001** |

**Figure 1.**
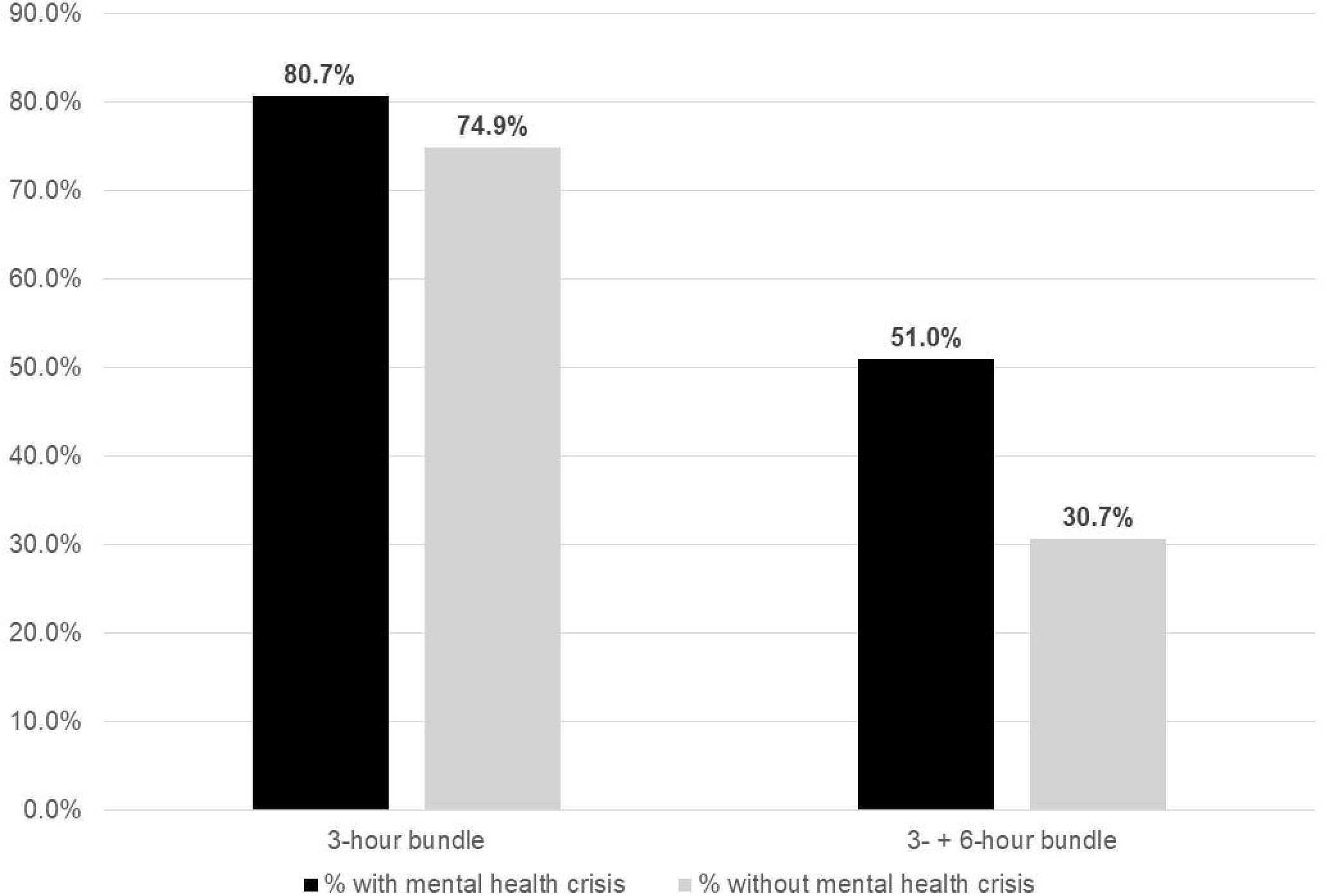

Among septic patients with a mental health crisis, there were no significant differences among those with cognitive (eg, dementia) vs. non-cognitive (eg, mood disorder) mental health crises with regards to 3- or 6-hour bundle care. (**Table 7, Figure 2**)

**Table 7.**

| Patient group | % receiving full sepsis bundle care | P-value |
| --- | --- | --- |
| 3-hour bundle, cognitive psychiatric disorder(s) | 76.1 | 0.3542 |
| 3-hour bundle, non-cognitive psychiatric disorder(s) | 86.7 |  |
| 6-hour bundle, cognitive psychiatric disorder(s) | 32.4 | 0.9437 |
| 6-hour bundle, non-cognitive psychiatric disorder(s) | 33.3 |  |
| 6-hour bundle, cognitive psychiatric disorder(s) | 42.6 | 0.7782 |
| 6-hour bundle, non-cognitive psychiatric disorder(s) | 38.5 |  |

**Figure 2.**
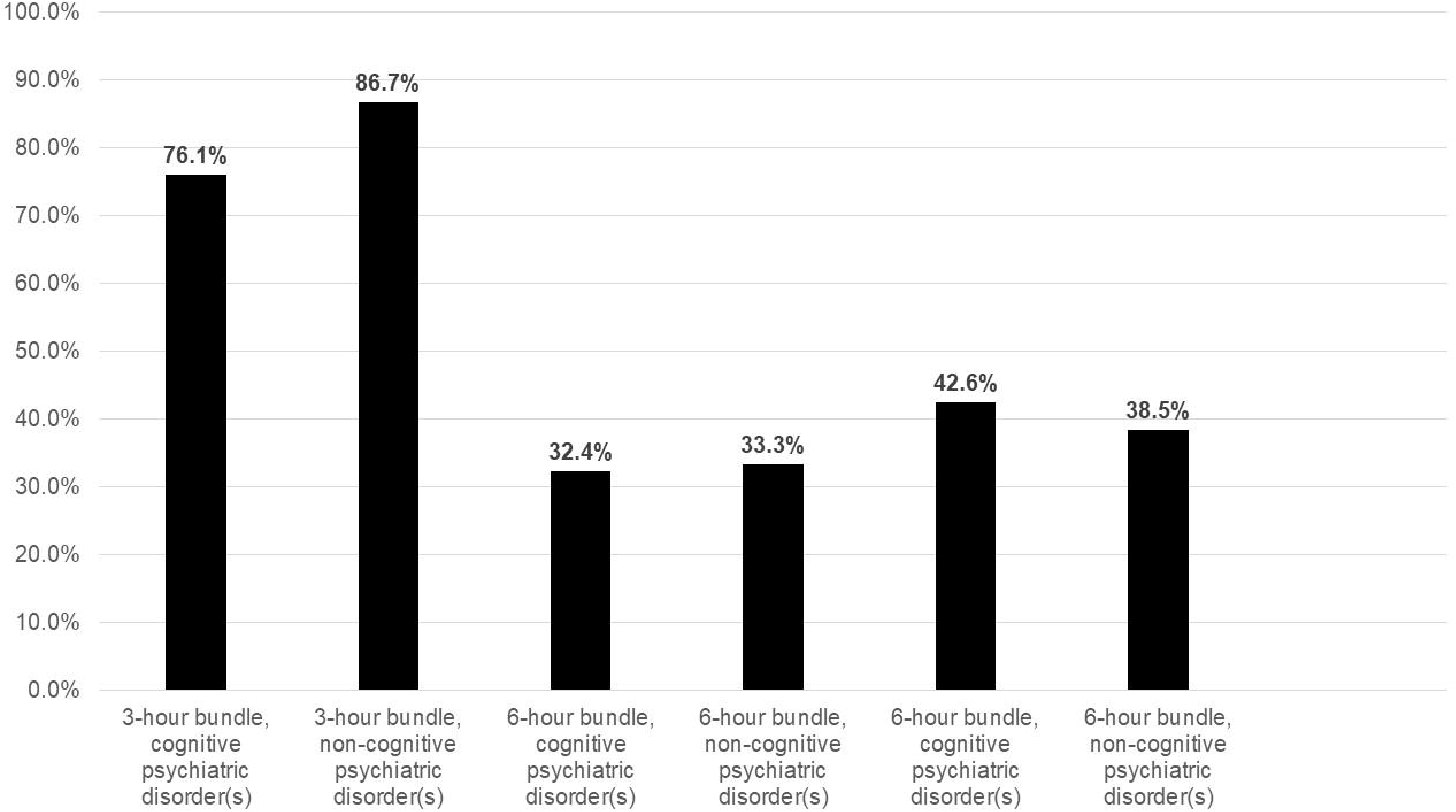

## Discussion

In this study, patients with and without an acute mental health crisis were equally-likely to receive full 3-hour Surviving Sepsis Campaign bundle care (∼80% vs. 75%, p = 0.1456). Among patients who received 3-hour bundle care, those with an acute mental health crisis were more-likely to receive 6-hour care (repeating serum lactate and re-assessing the need for additional IVF) (∼50% vs. ∼31%, p = 0.0001). This latter finding was contrary to our hypothesis, and in contradistinction to prior studies demonstrating patients with mental illness receive less-quality care than those without mental illness. We suspect this difference might be due to increased attention paid to patients with an acute mental health crisis, including more-frequent room visits by hospital staff or more concerns among family members. Among patients with an acute mental health crisis, there were no differences in the percent of patients with cognitive (eg, dementia) vs. non-cognitive (eg, psychosis, substance abuse) who received or did not receive full 3- or 6-hour sepsis bundle care. That is, no particular set of mental health conditions was associated with receiving or not receiving appropriate care.

### Limitations

This study has several limitations. First, we performed a univariate analysis, not a multivariate analysis. We did not adjust for potential confounding variables such as age, gender, comorbidities (immune suppression, CHF, CKD), current psychiatric medications, insurance type, or provider (MD or PA)) that might have explained the presence or absence of differences in sepsis care between patients with vs. without a mental health crisis. Future investigations of this dataset will include a logistic regression to control for such variables. In addition, we did not assess the impact of clinical variables, such as blood pressure, heart rate, lactic acid, and renal function on the aggressiveness of care, which could have influenced bundle compliance. For example, patients with poor renal function might not have received aggressive intravenous fluid (IV) resuscitation for fear of causing volume overload/congestive heart failure. Renal failure is more common among the elderly, and the elderly were more likely to have a mental health crisis (eg, dementia). Consequently, it’s possible those with dementia didn’t receive as much IV fluids, which could have lowered their bundle compliance rate. In such a circumstance, it wasn’t the dementia, but, rather, the renal dysfunction, that led to bundle non-compliance. Furthermore, this study did not investigate the impact of differing rates of 6-hour sepsis bundle compliance among those with vs. without a mental health crisis on sepsis outcomes such as mortality or ED, ICU, or hospital length-of-stay (LOS), which we intend to investigate in future studies.

## Conclusion

In contrast to our hypothesis, septic patients with and without an acute mental health crisis received similar quality of 3-hour sepsis bundle care. However, those with mental health crises received more-compliant 6-hour sepsis bundle care. That is, between hours 3 and 6, they were more-likely to have their lactic acid rechecked and response to IV fluids reassessed. This might be because such patients had more frequent visits by nurses or providers on account of their behavioral health issues, such as agitation.

## Data Availability

All data produced in the present study are available upon reasonable request to the authors

## Appendix A: ICD-10 codes for severe mental illness

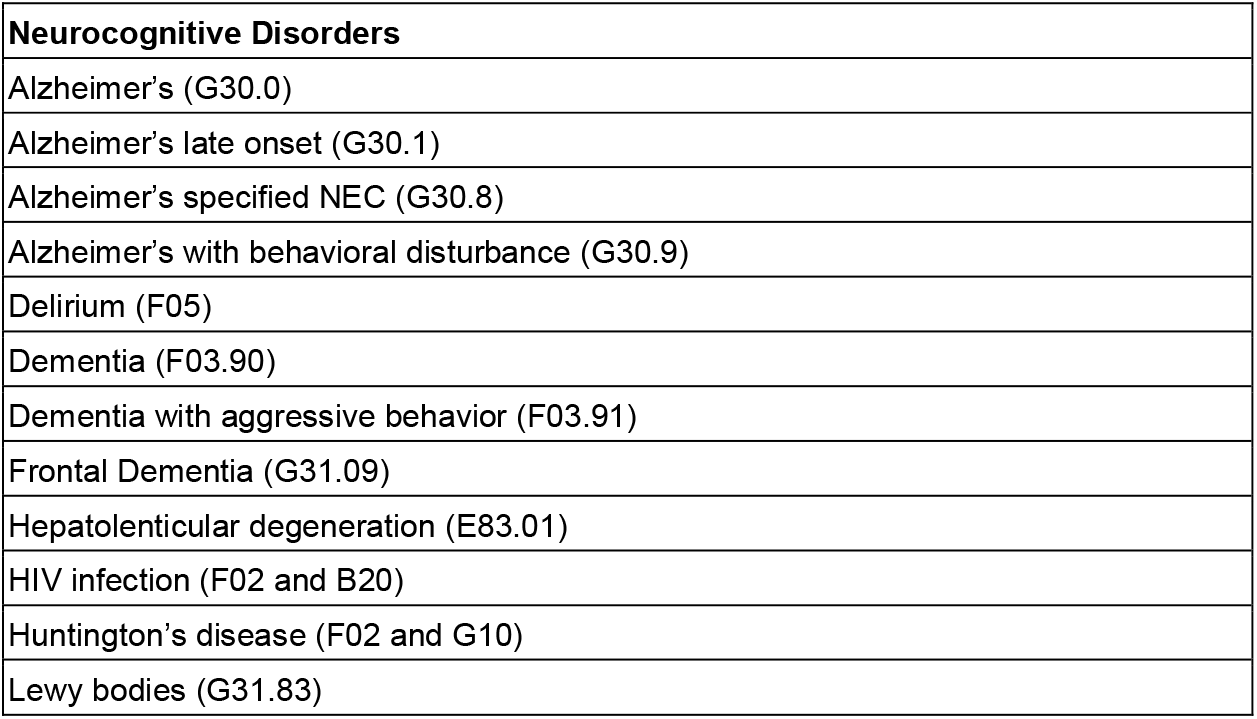
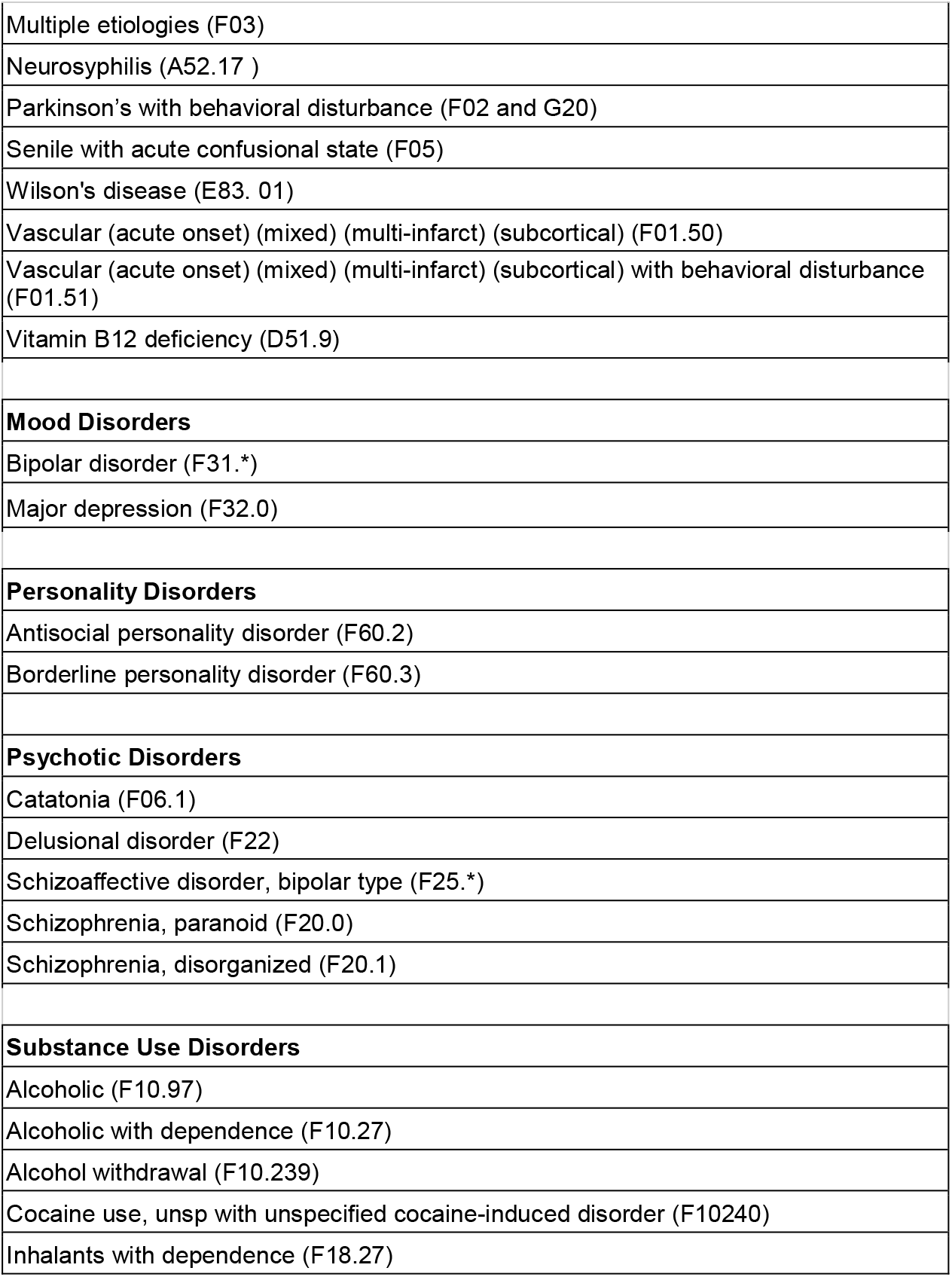
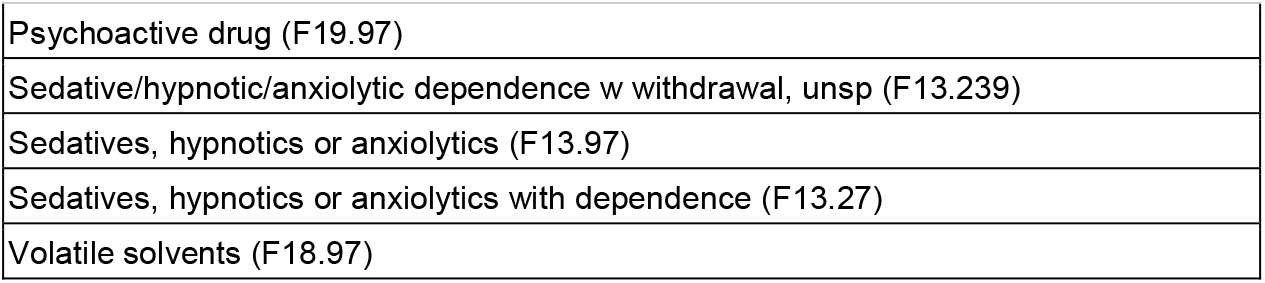

